# Comparison of BinaxNOW™ and SARS-CoV-2 qRT-PCR detection of the Omicron Variant from Matched Anterior Nares Swabs

**DOI:** 10.1101/2022.01.31.22270206

**Authors:** Lena Landaverde, Jacquelyn Turcinovic, Lynn Doucette-Stamm, Kevin Gonzales, Judy Platt, John H. Connor, Catherine Klapperich

## Abstract

The COVID-19 pandemic has increased the use of rapid antigen tests such as the Abbott BinaxNOW™ COVID-19 Antigen Self-Test. In winter of 2021-2022, the omicron variant surge made it quickly apparent that although rapid diagnostic tests (RDTs) are less sensitive than qRT-PCR, the accessibility, ease of use, and rapid read-outs of RDTs made them a sought after and often sold-out item at local suppliers. Here, we sought to qualify the BinaxNOW™ test for use in our university testing program as a method to rule-in positive or rule-out negative individuals quickly when they seek care at our priority qRT-PCR testing site. To perform this qualification study, we collected matched additional swabs from individuals attending this test site for standard of care qRT-PCR testing. All matched swabs were tested using the BinaxNOW™ RDT. Initially as part of a feasibility study, test period 1 (n=110) samples were put in cold storage prior to testing. In follow-on test period (n=209), we tested samples real-time at the test facility. Combined, 102 of 319 samples tested positive for SARS-CoV-2. All samples for which genome sequence could be collected were omicron (n=92). We observed a calculated sensitivity of 53.9%, specificity of 100%, a positive predictive value (PPV) of 100%, and a negative predictive value (NPV) of 82.2% for the BinaxNOW™ tests (n=319). Sensitivity improved (75.3%) by changing the qRT-PCR positivity threshold from a C_T_ of 40 to a C_T_ of 30. The ROC curve shows that for qRT-PCR positive C_T_ values between 24-40, the BinaxNOW™ test is of limited value diagnostically. Our results suggest that RDT tests could be used in our setting to confirm SARS-CoV-2 infection in individuals with substantial viral load, but that a significant fraction of infected individuals would be missed if we used RDT tests exclusively to rule out infection.

## Introduction

Experience in the winter of 2020 suggested that 2021 fall/winter holiday travel would also lead to an increase in SARS-CoV-2 positivity rates (1). This prediction was confirmed when holiday travel coupled with the emergence of the omicron variant (B.1.1.529) initiated unprecedented levels of infection beginning in December of 2021. Omicron accounted for most cases in the US a few weeks after it was first detected in the US on December 1, 2021 (2–4). The omicron surge overwhelmed many existing qRT-PCR diagnostic sites, driving an increased use of rapid diagnostic tests (RDTs). These tests rely on viral antigen detection and were developed to recognize SARS-CoV-2 variants that existed before the appearance of the highly mutated omicron lineage.

The Abbott BinaxNOW™ COVID-19 Antigen Self-Test (BinaxNOW™, Abbott, Des Plaines, IL) has been at the forefront of rapid diagnostic testing in the United States. However, debate about the effectiveness of these tests in different use cases has led to efforts to track their specificity and sensitivity as new variants emerge (5–10). At the time of this study, there is an omicron limit of detection (LoD) dilution study (11) and a selective study focused on reported lower limits of detection (LLOD) C_T_ value ranges for BinaxNOW™ (12). These studies suggested that there was analytical strength in the RDT, but also suggested a limited range of viral loads in which the assays consistently returned positive results from PCR positive samples. Two additional studies look at a broader range of SARS-CoV-2 RDT tests include a 15-day study of individuals with no symptoms comparing delta and omicron on 3 RDT tests (13) and a study testing previously PCR tested frozen samples on 5 different RDT tests (14).

In January and February of 2022, Boston University Clinical Testing Laboratory (BU CTL) was testing all members of the BU campus community at least once a week using qRT-PCR as reported previously (15,16). Symptomatic individuals, previously positive individuals scheduled for follow on testing, or those deemed close contacts can elect to get a test at any time and are directed to a special priority testing site at the BU Health Services Annex. As a continuing improvement exercise aimed at reducing cost and speeding turnaround time for testing at this site, we investigated whether the BinaxNOW™ test could be used effectively. The investigation focused on whether RDT use could provide a rapid rule-in, rule-out of SARS-CoV-2 infection in this population.

To assess the differential sensitivity for SARS-CoV-2 detection between our qRT-PCR test and BinaxNOW™ RDTs, we collected an additional swab from a series of individuals attending our test site described above. These extra swabs were tested using BinaxNOW™ tests. Two sets of samples for RDT testing were collected: the first set of additional swabs were collected, refrigerated, and were all tested at the end of the study period, while a second (and larger) set of additional swabs were tested on site within 15 minutes of collection. The results from these matched swabs were compared to qRT-PCR results. All positive samples were sequenced to determine the SARS-CoV-2 variant.

## Methods

### Sample Collection

Participants in the qRT-PCR testing program were asked to give a second swab in a sequential manner as they arrived at the BU Health Services Annex for their scheduled appointment. Each participant provided one additional anterior nares (AN) swab immediately after they provided their initial swab for the routine qRT-PCR test. Individuals swabbed both nostrils while observed by the on-site testing personnel. Individuals testing at this location can be symptomatic, a close contact of a positive individual, or a suspected positive individual coming for a confirmatory test. At this site, the qRT-PCR sample is collected using an ORAcollect•RNA swab (DNA Genotek, Inc., Ontario, Canada). The additional matched AN swab was taken using a Puritan Sterile Foam Tipped Applicator (Puritan, Guilford, Maine) swab that was placed into a dry, sterile 15 ml conical tube.

Qualification testing of the BinaxNOW™ RDT was first conducted on January 10^th^ and January 12^th^, 2022. In this study, 110 sets of paired AN swabs were collected from 106 individuals (4 individuals tested on both dates) as observed self-collections (110 ORAcollect•RNA for PCR and 110 Puritan for BinaxNOW™ tests). Samples taken for the RDT were stored at the end of each collection day at 4°C, and a subset were tested on January 13^th^ with the BinaxNOW™ RDT. Since this was a qualification study, we prioritized and preferentially tested the qRT-PCR positive samples and 5 select negatives on January 13^th^ and moved the rest of the qRT-PCR negative Puritan swabs to –80°C until they were processed on January 23^rd^.

Later, to ensure that storing samples overnight before testing with the BinaxNOW™ RDT did not affect the results, we did follow-on testing on February 7-11^th^ and February 14-17^th^, 2022. During this test period, samples were collected using the protocol above, but the BinaxNOW™ RDT tests were run in real-time within 15 minutes of sample collection at the BU Health Services Annex. In this round of testing, 209 sets of paired AN swabs were collected from 195 individuals (14 individuals tested more than once in this round) as observed self-collections.

Altogether, 319 sets of paired swabs were collected during the two test periods from 300 individuals (19 individuals tested more than once during the combined study periods).

We received clearance from the BU Charles River Campus IRB to publish the results, as the work was ruled not human subjects research because we were evaluating the performance of the test and not accessing personal health information from the individuals (BU CRC IRB exemption #6402X).

### qRT-PCR Testing

ORAcollect•RNA swabs were processed, extracted, and tested by qRT-PCR at the BU CTL as detailed in Landaverde, *et al*. (15). The BU CTL test detects N1 and N2 targets and RnaseP as a human RNA control. All qRT-PCR tests for this work were performed individually and none were pooled.

### Abbott BinaxNOW™ COVID-19 Antigen Self-Test

In the first test period, the additional matched AN swabs were stored as described above and subsequently tested using the manufacturer’s instructions. Briefly, 6 drops of the provided buffer were added, the swab was inserted into the BinaxNOW™ card and rotated 3 times clockwise, then the card was sealed with the integrated adhesive strip. After 15 minutes, the test was read from the results window, and a photograph of the result was taken as a record of the test. A positive control provided in the kit was run on a separate test card to confirm the validity of the test kit. In the second test period, the additional matched AN swabs were tested at the site according to the manufacturer instructions within 15 minutes of collection.

### SARS-CoV-2 Sequencing

Whole genome sequencing was performed on RNA extracted from all qRT-PCR positive samples using the excess discarded ORAcollect•RNA solution. Sequencing was performed using the Illumina COVIDSeq Assay (Illumina, San Diego, California) and sequenced on an Illumina NextSeq 500 (17). Full length genomes for each amplified sample were then assembled through alignment to the Wuhan-Hu-1 reference sequence (NC_045512.2) (18) using Bowtie2(19). Nucleotide substitutions, insertions, and deletions were identified with LoFreq (20) Lineage assignment for each genome was carried out using Pangolin(21).

### Data Analysis

Data analysis was performed in MATLAB (MathWorks, Natick, Massachusetts). Sensitivity, specificity, positive predictive value (PPV), and negative predictive value (NPV) were determined for the BinaxNOW™ RDT.

## Results and Discussion

In the first test period (January 10^th^ and January 12^th^, 2022), 48/110 matched swabs were positive for COVID-19 by the standard of care qRT-PCR test. The C_T_ values for the positive samples ranged between 11.7-38.8 for N1 and 11.7-38.8 for N2 (**Figure 1A**). Using the BU CTL qRT-PCR test as the gold standard, the BinaxNOW™ tests detected 25/48 positive results, and 62/62 of the negative results (**Table 1, Supplemental Figure 1**), leading to a calculated sensitivity of 52.1%, specificity of 100%, a positive predictive value (PPV) of 100%, and a negative predictive value (NPV) of 72.9%.

**Table 1.** 2×2 Table for qRT-PCR and BinaxNOW™ matched samples.

| BU Clinical Testing Lab Classification |  |  |  |
| --- | --- | --- | --- |
| Test Period 1 (RDT run after swabs were in cold storage) |  |  |  |
|  | C<sub>t</sub> <40 | C<sub>t</sub> >40 |  |
|  | RT-PCR + | RT-PCR - | Totals |
| Binax + | 25 | 0 | 25 |
| Binax - | 23 | 62 | 85 |
| Totals | 48 | 62 | 110 |
| Test Period 2 (RDT run in real-time) |  |  |  |
|  | C<sub>t</sub> <40 | C<sub>t</sub> >40 |  |
|  | RT-PCR + | RT-PCR - | Totals |
| Binax + | 30 | 0 | 30 |
| Binax - | 24 | 155 | 179 |
| Totals | 54 | 155 | 209 |
| Combined |  |  |  |
|  | C<sub>t</sub> <40 | C<sub>t</sub> >40 |  |
|  | RT-PCR + | RT-PCR - | Totals |
| Binax + | 55 | 0 | 55 |
| Binax - | 47 | 217 | 264 |
| Totals | 102 | 217 | 319 |

**Figure 1.**
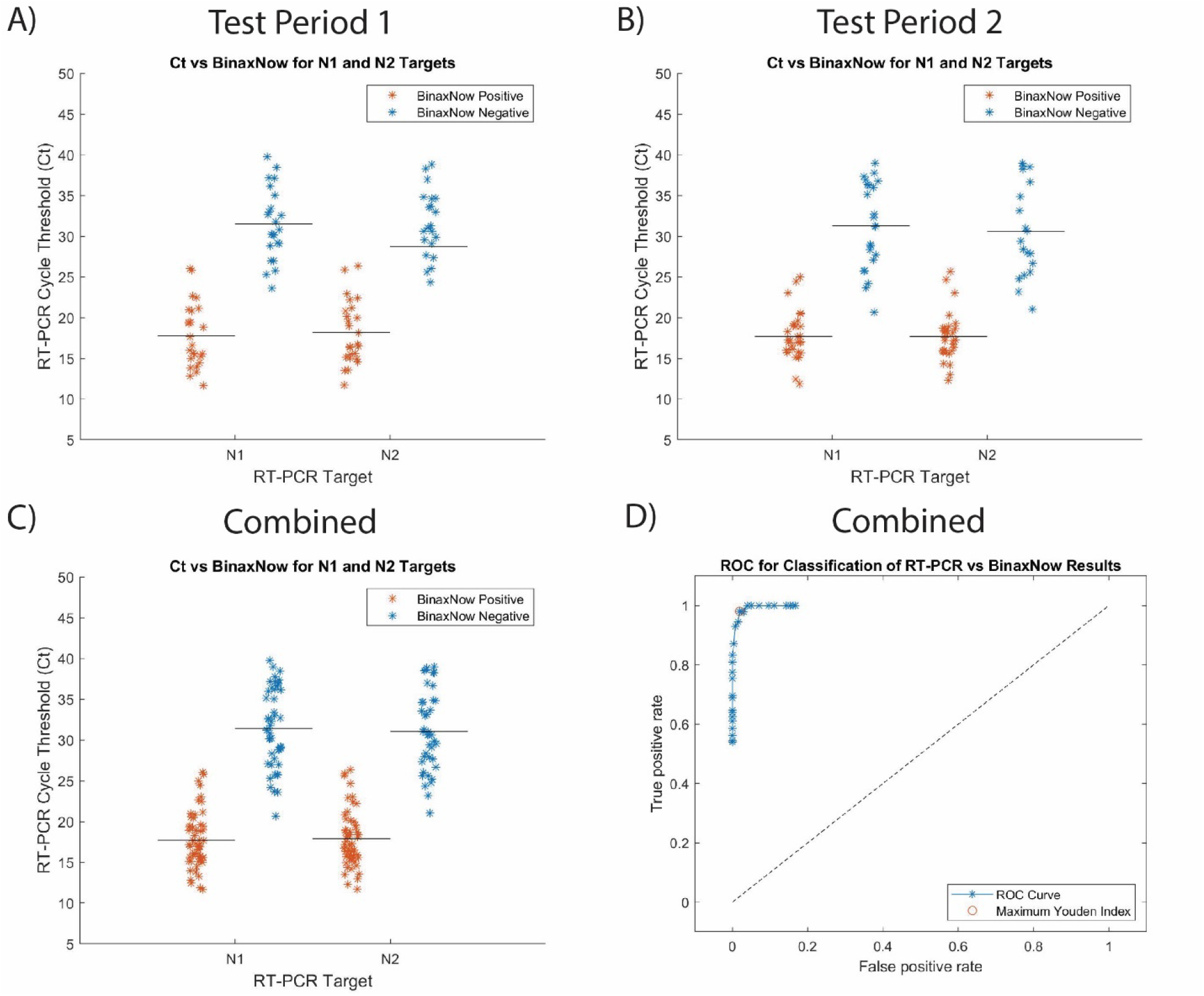
Plots showing the qRT-PCR positive sample C_T_ values by Target. **A)** In Test Period 1, the BinaxNOW™ RDTs were positive for C_T_ values from 11.7-26.0 for N1 and 11.7-26.3 for N2. The BinaxNOW™ RDT negative tests had C_T_ values from 23.6-39.8 for N1 and 24.3-38.8 for N2. The average C_T_ is 24.3 for N1 with a standard deviation of 8.0. The average C_T_ is 24.2 for N2 with a standard deviation of 7.7. **B)** In Test Period 2, the BinaxNOW™ RDTs were positive for C_T_ values from 11.9-24.9 for N1 and 12.3-25.6 for N2. The BinaxNOW™ RDT negative tests had C_T_ values from 20.7-39.0 for N1 and 21.0-39.0 for N2. The average C_T_ is 23.6 for N1 with a standard deviation of 7.9. The average C_T_ is 22.7 for N2 with a standard deviation of 7.5. **C)** The combined data set has an average C_T_ is 23.9 for N1 with a standard deviation of 7.9. The average C_T_ is 23.4 for N2 with a standard deviation of 7.6. **D)** ROC Curve for qRT-PCR vs BinaxNOW™ of the combined dataset using qRT-PCR C_T_ values ≤40 as the gold standard. Within each qRT-PCR target N1 and N2, data is summarized in Supplementary Table 1 for A, B, and C.

In the second test period (February 7-11^th^ and February 14-17^th^, 2022), 54/209 matched swabs were positive for COVID-19 by the qRT-PCR test. The C_T_ values for the positive samples were between 11.8-39.0 for N1 and 12.2-39.0 for N2 (**Figure 1B**). This resulted in a calculated sensitivity of 55.6%, specificity of 100%, a PPV of 100% and an NPV of 86.6% (**Table 1, Supplemental Figure 1)**.

A total of 100 positive samples had enough residual material after testing for sequencing, 92/100 were successfully sequenced and 100% (n=92) were the omicron variant.

The mean and standard deviation were similar for both test periods (**Supplementary Table 1**). Statistically there is no difference between the test periods for the N1 (Kruskal-Wallis test). There was a small but significant difference between the two sample collection groups for the N2 target. The combined data set from the two study periods yielded a sensitivity of 53.9%, specificity of 100%, PPV of 100%, and a NPV of 82.2% (**Table 2**). The combined C_T_ values for positive samples range between 11.6-39.8 N1 and 11.7-39.0 N2 (**Figure 1C**).

**Table 2.** Table for combined data set sensitivity with various C_t_ cutoffs (5)

|  |  | BU CTL Classification | Pilarowski, et al Classification | ROC Curve |
| --- | --- | --- | --- | --- |
|  | All Data | C<sub>t</sub> ≤ 40 | C<sub>t</sub> ≤ 30 | C<sub>t</sub> ≤ 24 |
| Sensitivity | 53.9 | 53.9 | 75.3 | 94.8 |

The sensitivity calculations depend heavily on established criteria for positivity. The cut off for a positive result for the BU CTL qRT-PCR test is a C_T_ ≤40 for one or both of the N1 and N2 targets. Other studies have advocated for using a modified C_T_ value that reasoning that individuals who have higher C_T_ values may not transmit virus (22,23). For example, Pilarowski, *et al*. use a C_T_ cutoff of 30 for positivity (**Supplemental Table 2**). (5). Using that cutoff for our combined dataset, our revised sensitivity would be 75.3%, specificity 100%, PPV 100%, and NPV 93.2% (**Table 2**). In either case, the sensitivity is significantly lower than the initially published 93.3% for BinaxNOW™ (5).

Here, the BinaxNOW™ RDT returned variable test results at C_T_ values between 20.7-26.0 for N1 and 21.0-26.3 for N2 (**Figure 1C**). The mean C_T_ value is 17.7 with a standard deviation of 3.4 for N1 and mean C_T_ value is 17.9 with a standard deviation of 3.4 for N2 when both qRT-PCR and BinaxNOW™ are positive (**Supplementary Table 1**). From the data, samples with a C_T_ lower than 24 are highly likely to test positive on the BinaxNOW™ RDT. These results are consistent with a smaller study on an omicron outbreak that paired saliva samples tested with RT-PCR and nasal rapid antigen tests (Quidel QuickVue™ At-Home OTC COVID-19 Test and Abbott BinaxNOW™ COVID-19 Antigen Self-Test)(24).

**Figure 1D** shows an ROC curve constructed the full data set. The maximum Youden index for the ROC curve identifies where both sensitivity and specificity are at maximum (25,26). The Youden Index of 0.96 corresponds to a C_T_ of 24 for the qRT-PCR cycle cutoff (**Figure 1D**). This result provided important information regarding our ability to implement BinaxNOW™, antigen-based testing, as a surrogate for qRT-PCR testing. A cutoff value of 24 for qRT-PCR is below the average C_T_ value reported for both delta and omicron variants (5,6,8,9,12,13), suggesting that RDT testing alone would be insufficient to maintain control of spread in our community (**Supplementary Table 3**). Thus, the conclusion of our qualification study was that it is not possible to use the BinaxNOW™ test alone to replace qRT-PCR in our testing sites for ruling-out positive individuals. We estimate that had we only used the RDT, we would have missed 46% of the cases.

It was already known, and expected, that RDTs, like BinaxNOW™, have a lower sensitivity compared to qRT-PCR for detecting SARS-CoV-2 (5–10). The data presented here extends this conclusion to include a real-world application where almost all the cases were confirmed to be the omicron variant. As more data emerges linking transmission rates with viral loads, more solid conclusions will be made about the best use cases for RDTs. Until that time, it is still prudent to be cautious about a negative BinaxNOW™ test, especially when symptoms are present.

Limitations of this study include the relatively small sample size compared to other studies of the BinaxNOW™ RDT earlier in the pandemic (6). The disease prevalence at our clinic measured by qRT-PCR on the first two days of testing was approaching 50%, and for the second round of testing was about 25%. Recall that this site was biased toward symptomatic individuals and close contacts of known cases. The mean prevalence during the first test period in our entire population (approximately 40,000 people) was 5.0% and in the second test period it was 0.9%.

It is also important to note that we asked individuals to swab each nostril twice, and the swab for the BinaxNOW™ test was always taken second. It is possible that less material was present on the swabs taken for the RDT, which could account for some of the lower sensitivity. Finally, we would like to note that 19 individuals tested more than once and thus, our combined dataset includes 319 matched samples from 300 individuals. We do know that the 19 individuals who tested two or more times never tested positive more than once.

To be sure, tests like BinaxNOW™ are valuable tools, as they provide immediate results, require no instrumentation, and are highly effective at rule-in diagnosis. However, there is still an unmet need for more sensitive rapid diagnostics for SARS-CoV-2 that could augment qRT-PCR testing at times of high demand.

## Data Availability

All data produced in the present study are available upon reasonable request to the authors

## Supplementary Information

**Supplementary Figure 1.**
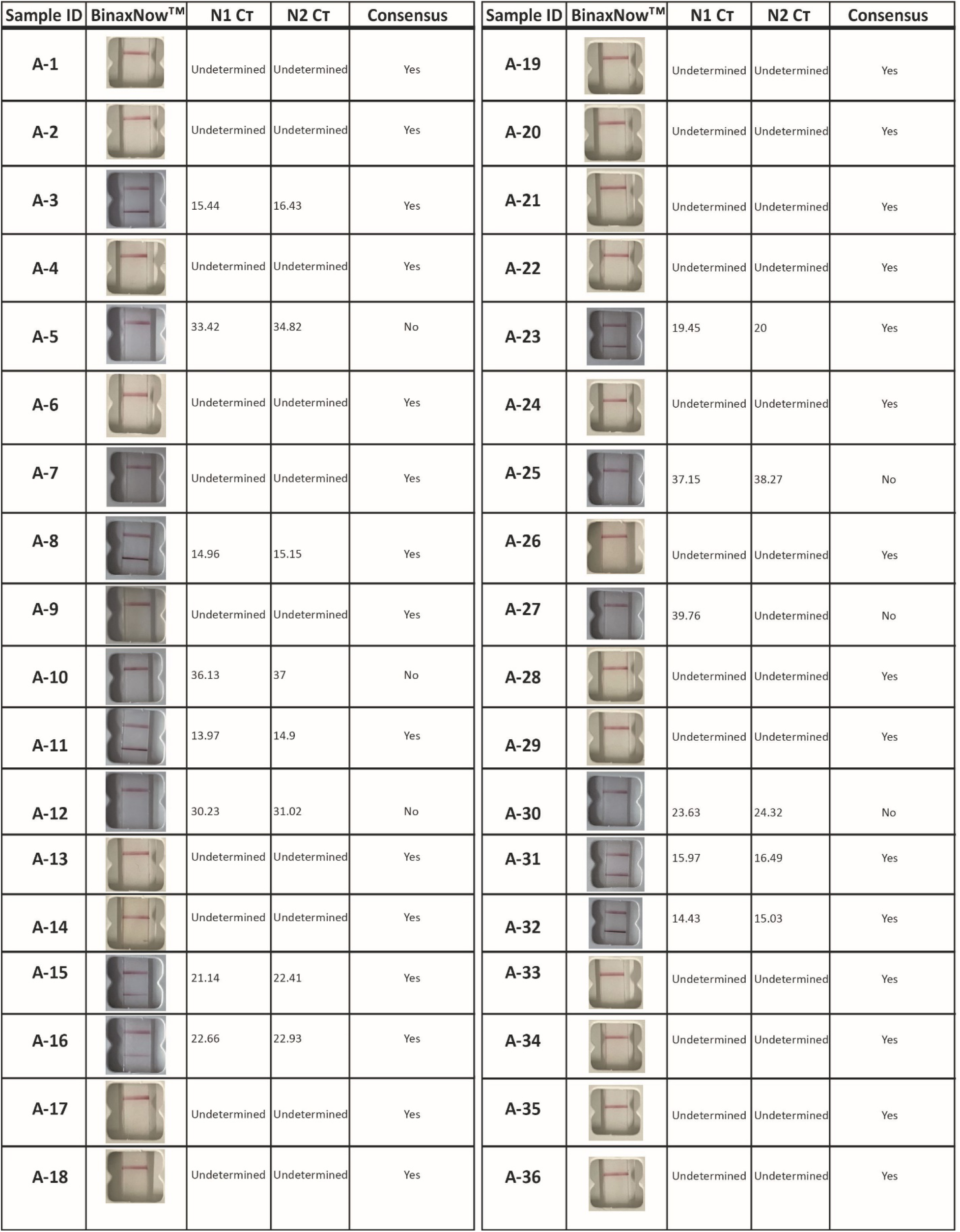
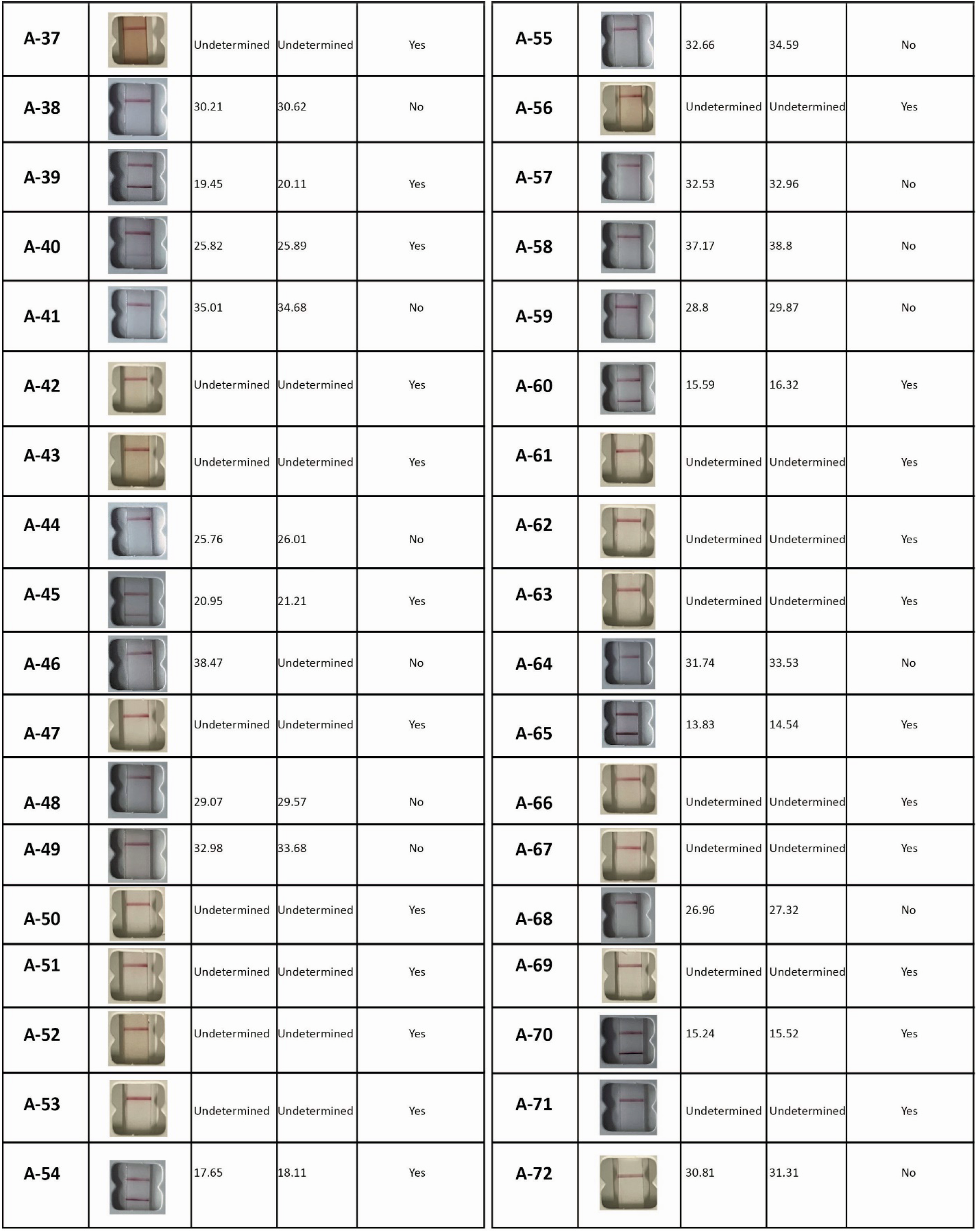
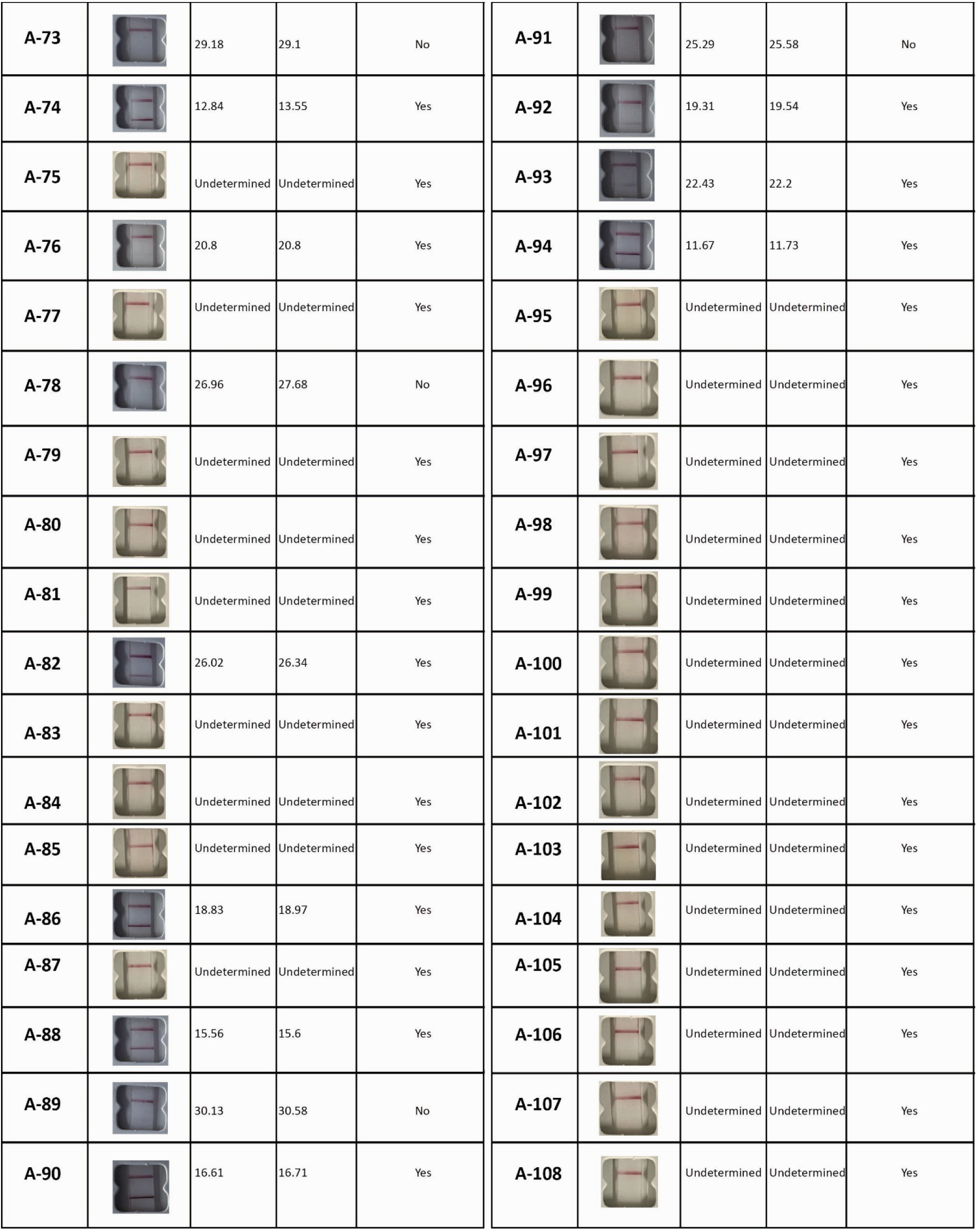
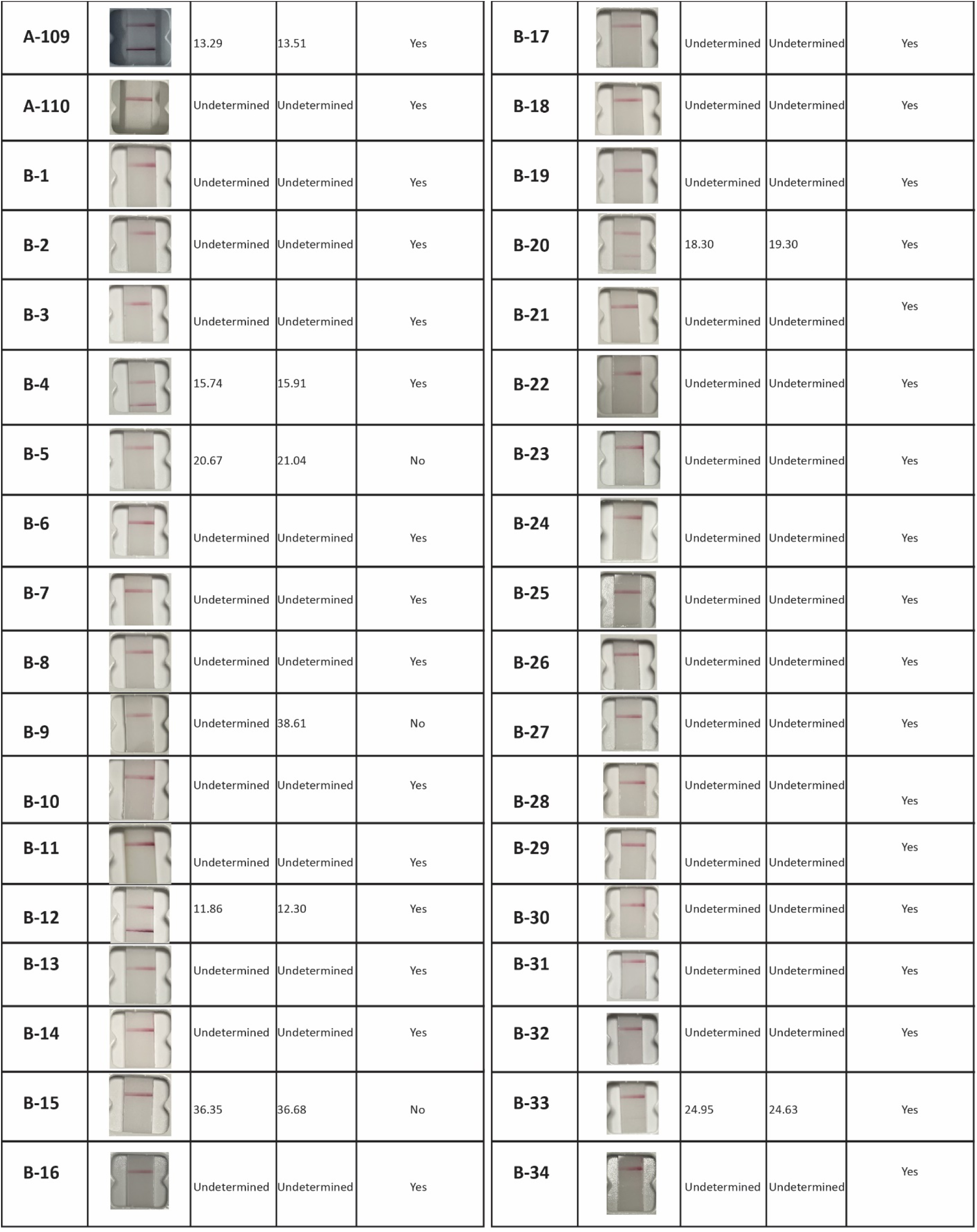
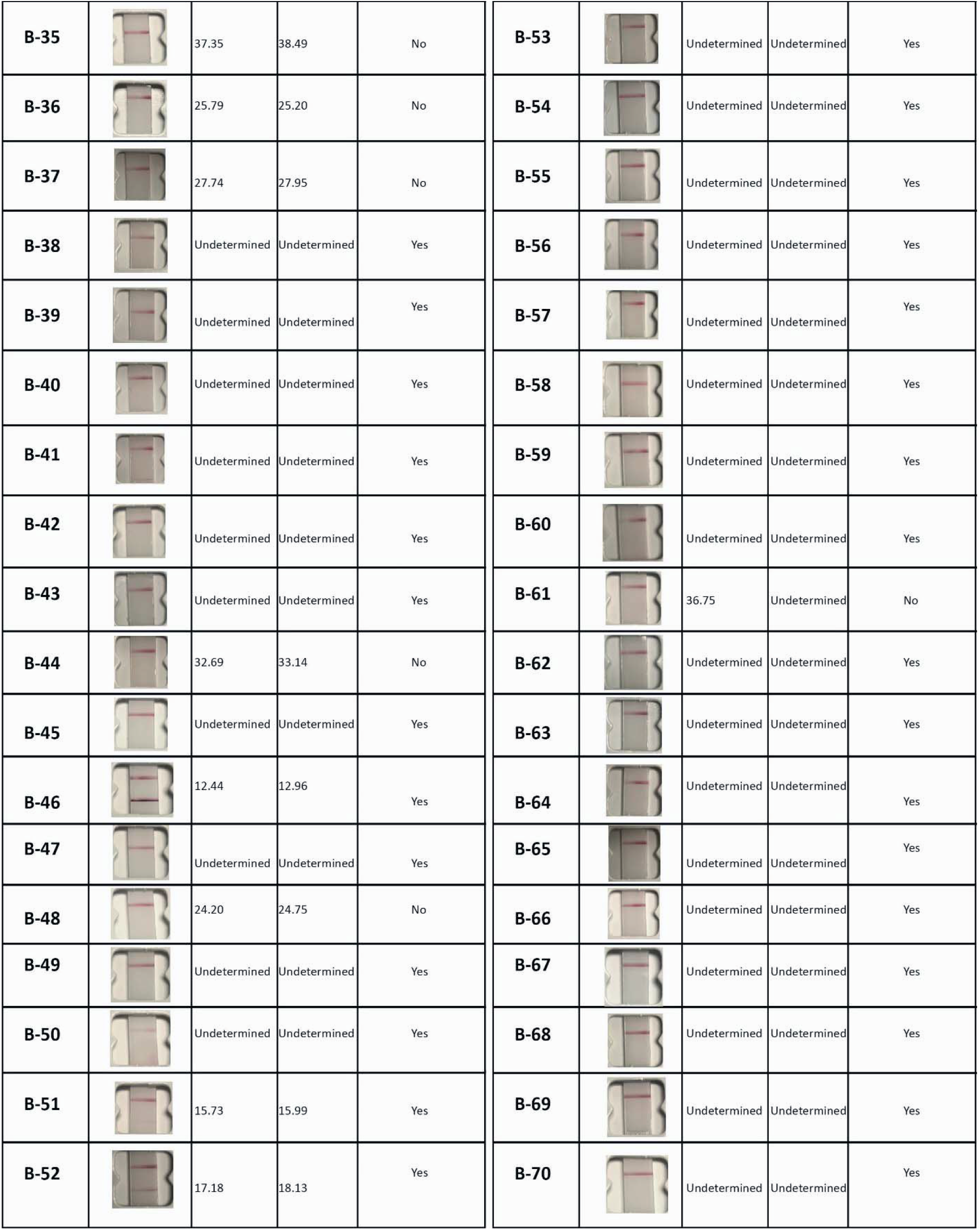
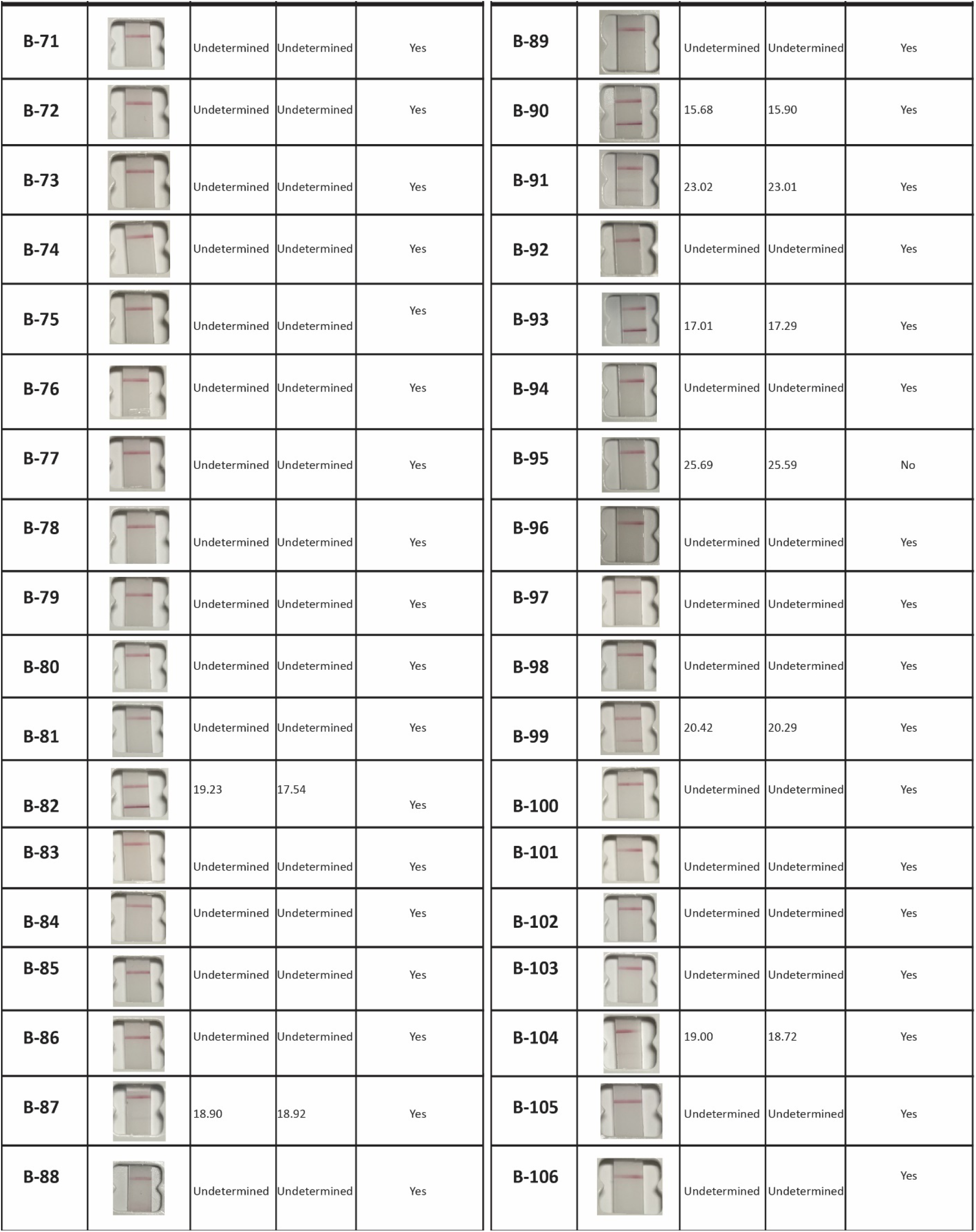
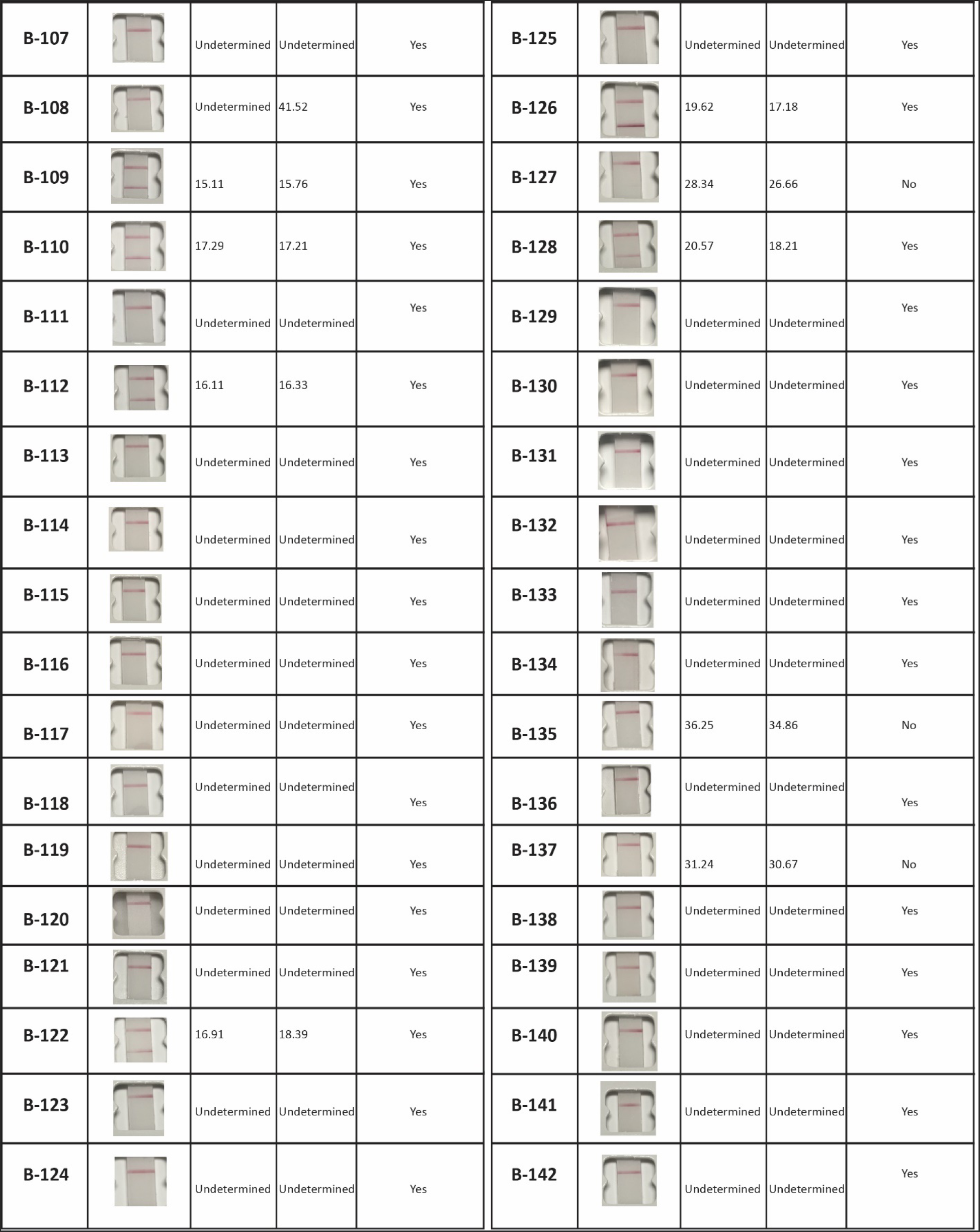
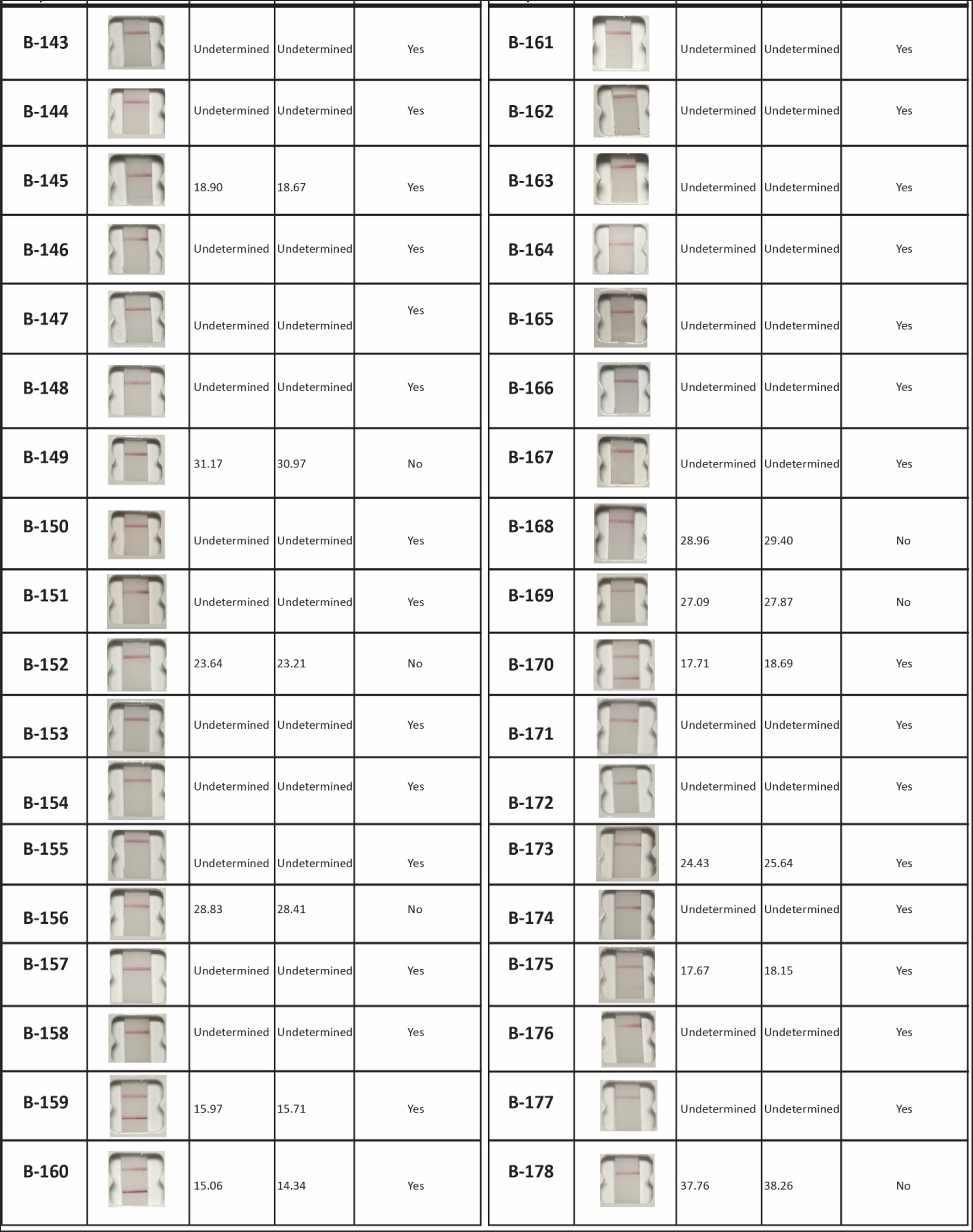
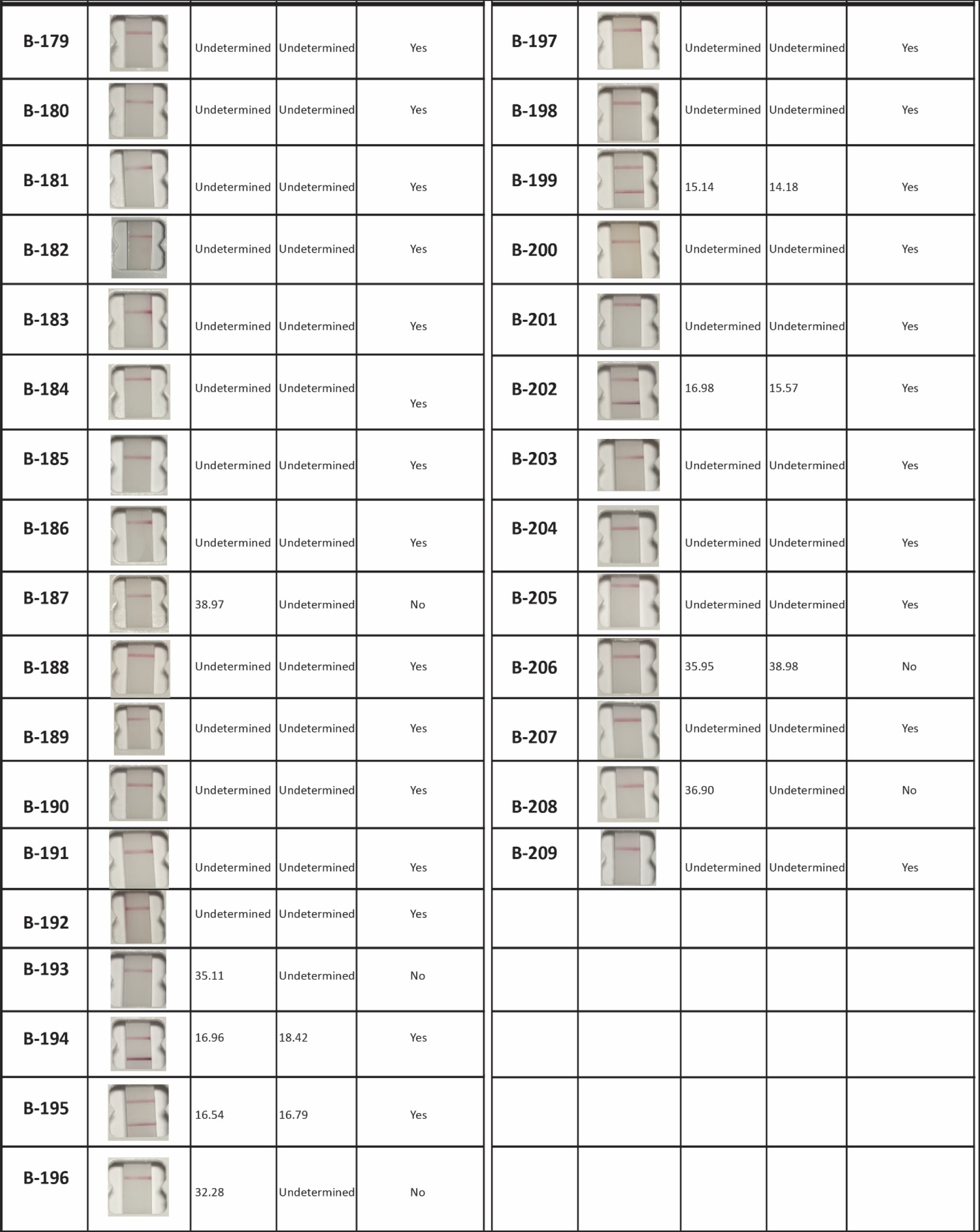
Summary of data set with qRT-PCR and BinaxNOW™ RDT. Photographs of all BinaxNOW™ test cards run in this study. Tests A-1 through A-110 were performed on swabs that had been kept in cold storage for one or more days. Tests B-1 through B-209 were run immediately after swabbing at the test site. None of the images have been altered or adjusted, since these were taken only as a record of each test. They were taken with an iPhone, 15 minutes after the test was started.

**Supplementary Table 1.**
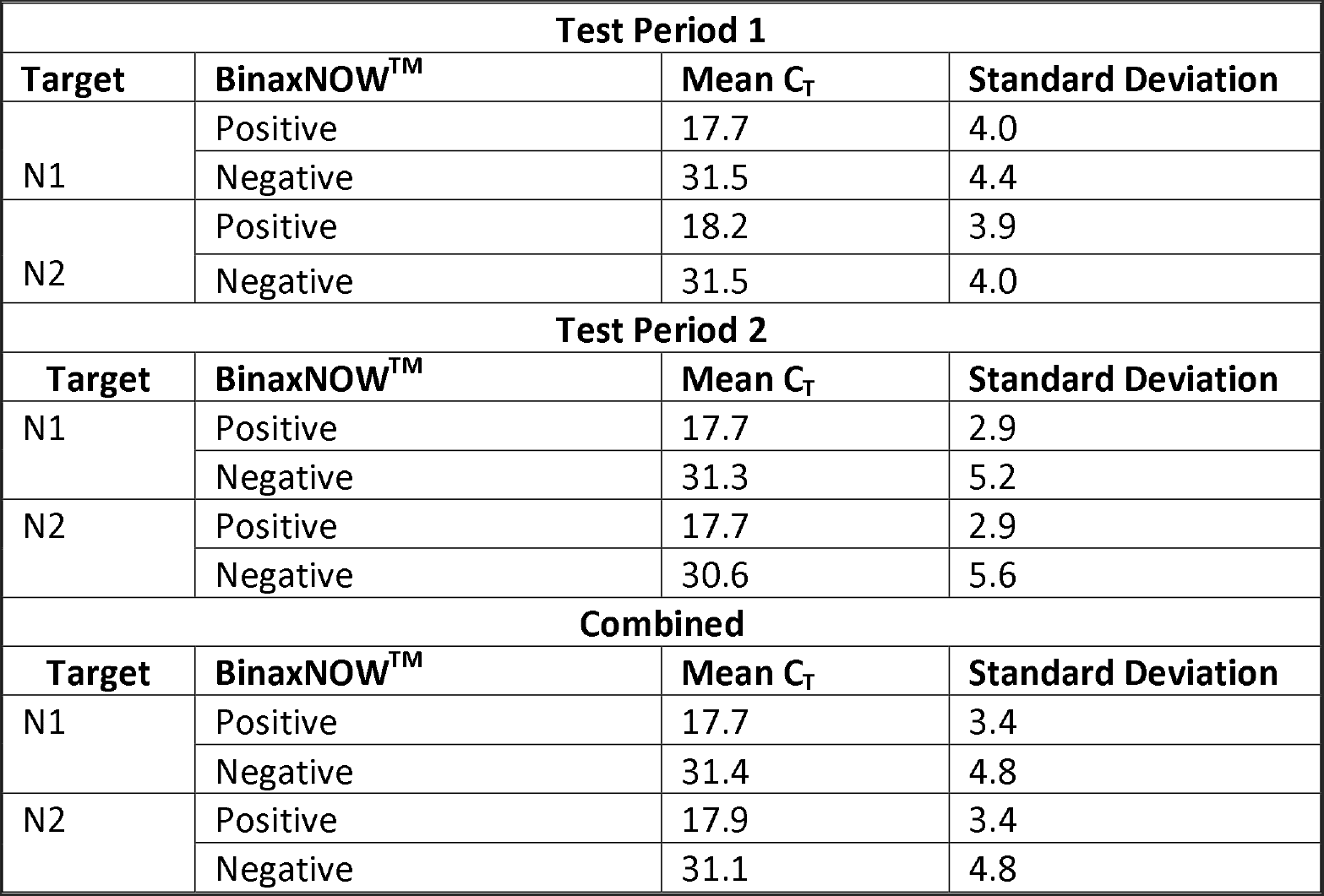
Mean and Standard Deviation of qRT-PCR positive by BinaxNOW™ result and targets.

| Test Period 1 |  |  |  |
| --- | --- | --- | --- |
| Target | BinaxNOW™ | Mean C<sub>T</sub> | Standard Deviation |
| N1 | Positive | 17.7 | 4.0 |
|  | Negative | 31.5 | 4.4 |
| N2 | Positive | 18.2 | 3.9 |
|  | Negative | 31.5 | 4.0 |
| Test Period 2 |  |  |  |
| Target | BinaxNOW™ | Mean C<sub>T</sub> | Standard Deviation |
| N1 | Positive | 17.7 | 2.9 |
|  | Negative | 31.3 | 5.2 |
| N2 | Positive | 17.7 | 2.9 |
|  | Negative | 30.6 | 5.6 |
| Combined |  |  |  |
| Target | BinaxNOW™ | Mean C<sub>T</sub> | Standard Deviation |
| N1 | Positive | 17.7 | 3.4 |
|  | Negative | 31.4 | 4.8 |
| N2 | Positive | 17.9 | 3.4 |
|  | Negative | 31.1 | 4.8 |

**Supplementary Table 2.** 2×2 Table for qRT-PCR and BinaxNOW™ matched samples using the Pilarowski, *et al*. classification (5).

| Pilarowski, *et al.* Classification |  |  |  |
| --- | --- | --- | --- |
| Combined |  |  |  |
|  | C<sub>t</sub> <30 | C<sub>t</sub> >30 |  |
|  | RT-PCR + | RT-PCR - | Totals |
| Binax + | 55 | 0 | 55 |
| Binax - | 18 | 246 | 264 |
| Totals | 73 | 246 | 319 |

**Supplementary Table 3.** 2×2 Table for qRT-PCR and BinaxNOW™ combined matched samples with maximum Youden Index classification (25,26).

| **ROC Curve Maximum Youden Index (0.98) Classification** |  |  |  |
| --- | --- | --- | --- |
| **Combined** |  |  |  |
|  | C<sub>t</sub> <24 | C<sub>t</sub> >24 |  |
|  | **RT-PCR +** | **RT-PCR -** | Totals |
| **Binax +** | 51 | 4 | 55 |
| **Binax -** | 3 | 261 | 24 |
| Totals | 54 | 265 | 319 |

